# Exploring The Lived Experiences Of Depression Among Youths And Their Guardians In Sub-Saharan Africa: A Systematic Review and Meta-Aggregation

**DOI:** 10.1101/2025.05.06.25327134

**Authors:** Precious Makiyi, Wisdom Malata, Michael Udedi, Moses Kumwenda, Adamson Muula, James January

## Abstract

**Background:** The cultural context influences how mental disorders like depression, are perceived and experienced. Most studies on lived experiences of depression by youths have been done in high-income countries. This has made it difficult for researchers to infer the findings in the context of SSA due to differences in cultures and socio-demographic characteristics. This systematic review thus, seeks to understand how depression is experienced by youths and their guardians in SSA.

**Objective:** This systematic review was done to address two key objectives: to explore and synthesize qualitative studies on the lived experience of depression among youths and their guardians and to identify common themes, and the impact of socio-cultural and contextual factors on the lived experience of depression among youths and their guardians in SSA.

**Methodology:** Two research assistants independently searched and selected articles from databases such as PubMed, using a set search strategy. Thereafter, two reviewers reviewed and extracted data from the selected articles, analysed it using meta-aggregation techniques. The identified papers underwent an independent quality assessment using the Critical Appraisal Skills Program checklist by the two independent researchers. Through this quality assessment, the risk of bias in the papers was also assessed. We have used the Preferred Reporting Items of Systematic Reviews and Meta-analyses (PRISMA) guidelines in reporting this review.

**Results:** Our search resulted into 14 unique studies spanning 8 countries in SSA, representing a total of 610 young people (10 to 24 years; 72% females) with lived experience of depression, 297 caregivers and 70 healthcare workers. Findings on the lived experience of youths were aggregated into eleven categories which were further grouped into three main synthesised findings of making sense of depression, mental health systems and services and contextual issues as key issues encompassing their lived experience. Regarding the experience of guardians, three key themes were identified: challenges faced by the guardians, their perception of youth depression and their experience in navigating through the healthcare system.

**Conclusion:** the sociocultural context in SSA determines how the youth experience depression and that symptoms of depression as experiences by youths in SSA may not always match diagnostic criteria developed from the west. The experiences of the guardians have an impact on the young people’s experiences of depression.

**Systematic review registration**: PROSPERO, CRD42024556661

## Background

Depression among adolescents and young people, is one of the major public health issues of concern globally and Sub-Saharan Africa, in particular (1). Studies indicate that 50% of adult mental health disorders start by age 14 and that about 75% of such cases start before 24 years of age (1). A Systematic Review on the prevalence of mental health problems among sub-Saharan African adolescents which included nine studies involving 14,409 adolescents in the general population found a median point prevalence for depression of about 27% (2), lesser than an average global rate of 34% (3). A distribution of the global cases, however, indicate that the majority are in Africa, Middle East and Asia. A comparison of global trend of point prevalences of depressive symptoms among adolescents between 2001 -2010 period and 2011-2020, showed an increase from 24% to 37% (3). There are associated predictions that the rates of depression among the youth and adolescents were more likely to increase due to various global challenges including the recent COVID-19 pandemic (4).

In SSA, context-specific risk factors such as poverty, HIV and early and teen pregnancies predispose the youth to depression (5,6). A systematic review on the prevalence of common mental disorders among young people living with HIV (YPLWH) in SSA found rates of between 16% and 40% for major depression and 4.4% to 52% for depressive symptoms (regardless of the severity) (7)

Studies done across the globe to better understand how the youth experience depression (1,8), show that despite the biological, psychological, sociological and cognitive differences between adolescents and adults, both the Diagnostic and Statistical Manual of Mental Disorders Version 5 (DSM-5) (9) and the International Classification of Diseases-11 (ICD-11) (10) do not have a proper distinction between adult and adolescent/youth depression save for the DSM-5 which only recognises irritability as an alternative to low mood (11). However, in practice, youth depression and adult depression present differently with the former depicting unique features (11).

Other studies indicate that, theories on adolescent depression are mainly based on adult depression, ignoring the influence of the neuro-developmental stages of adolescence (11). Despite the high risk of developing mental disorders as discussed above, young people’s access to mental health support is limited and the support services available to them are usually fragmented, non-encompassing, non-flexible and not youth-friendly (1). This has had a bearing on seeking mental healthcare among the youths and on untreated depression cases and related consequences which include but not limited to poor work and academic performance, increased indulgence in risky behaviours like use of psychoactive substances, loss of self-worth, family conflicts, reduced social functioning and increased risk of suicidal behaviours (12,13).

Against the synopsis of the epidemiology of youth depression as discussed above, it is imperative to get an understanding of how the youths in SSA and their guardians, experience depression. Unlike in the adult population, where lived experiences of depression are comparatively well-documented, little is known about the lived experience of depression by the youth, especially those in SSA (13). The advantage of exploring the lived experience of depression by the youth themselves is that their perspective provides a valuable information which is crucial in the development of theories on youth depression as well as youth-friendly interventions (8).

### Objectives of the Systematic review and Meta-aggregation

This systematic review and meta-aggregation have been conducted to address the following specific objectives:

- To understand the first-hand experience of developing and living with depression by youths and their guardians in SSA
- To determine the impact of contextual factors on the lived experience of depression among adolescents and their guardians in SSA
- To critically appraise the qualitative studies on lived experience of depression by the youth in SSA.

## Methodology

The review was registered with [PROSPERO](https://www.crd.york.ac.uk/PROSPERO/view/CRD42024556661) (CRD42024556661), where a brief copy of the protocol can be accessed (14). Additionally, the full protocol for the review was submitted to a journal and is under peer review. The systematic review is being reported in accordance with the Preferred Reporting Items for Systematic Reviews and Meta-Analyses (PRISMA) guidelines (15).

### Information Sources

The following electronic databases were used to search for related papers to our systematic review question: PubMed, Embase, Google Scholar, CINAHL and Scopus. The databases were searched from inception until August 30, 2024. We only considered those written in |English. Where applicable, we contacted authors of included reviews for clarification on their published information as well as to get additional information. We included one grey literature in the form of a conference abstract to enrich our data. Additionally, we examined reference lists of related articles to identify relevant journal papers to include in the review. We also did forward citation searches with no success.

### Search Strategy

The Following search concepts were included in our search strategy: 1. Youth or Adolescents or caregivers 2. Depression, 3. Phenomenology 4. Perception or experience and 5. Sub-Saharan Africa. The above-mentioned databases had tailored controlled vocabulary words. Figure 1 illustrates an electronic search strategy that was used in one of the databases (PubMed). Our search terms and subsequently, strategy as used in PubMed is as follows:

**Fig 1:**
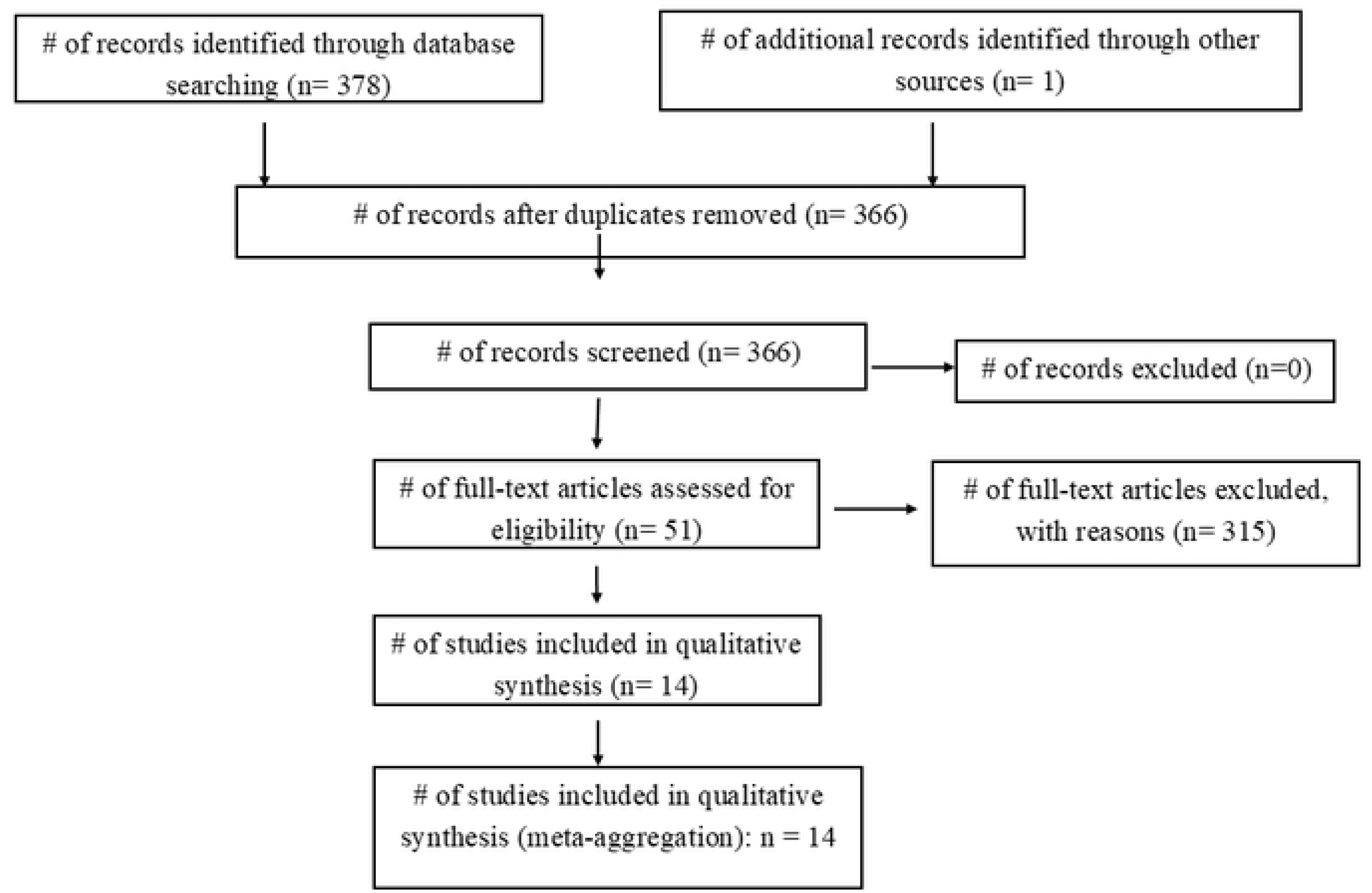
PRISMA Flowchart of study selection process.

### Eligibility Criteria

Table 2 shows the inclusion and exclusion criteria that was used in identifying the selected articles. The criteria were based on the modified version of the Population, Intervention, Control and Outcomes (PICO) Format.

**Table 1:** Search Strategy.

| Database | Search Strategy |
| --- | --- |
| PubMed | (((((Adolescent[MeSH Terms] OR adolescen*[MeSH Terms] OR teen*[MeSH Terms] OR youth[MeSH Terms] OR guardian[MeSH Terms] OR parent*[MeSH Terms] OR carer*[MeSH Terms] OR caregiver[MeSH Terms])) AND (Depression[MeSH Terms] OR Depressive disorder[MeSH Terms] OR Irritable mood[MeSH Terms] OR Anhedonia[MeSH Terms] OR mood disorders[MeSH Terms] OR Depress*[MeSH Terms] OR Dysthymi*[MeSH Terms] OR mood disorder*[MeSH Terms] OR affective symptom*[MeSH Terms] OR affective disorder*[MeSH Terms] OR melanchol*[MeSH Terms] OR psychological distress*[MeSH Terms] OR Irritable mood*[MeSH Terms])) AND (focus group[MeSH Terms] OR grounded theory[MeSH Terms] OR interviews[MeSH Terms] OR ethnograph*[MeSH Terms] OR hermeneut*[MeSH Terms] OR autobiography[MeSH Terms] OR biograph*[MeSH Terms])) AND (Perceived[MeSH Terms] OR beliefs[MeSH Terms] OR narrative[MeSH Terms])) AND (Africa[MeSH Terms] OR Sub-Saharan Africa[MeSH Terms] OR low-income[MeSH Terms] OR tropic*[MeSH Terms] OR afro* OR Angola OR Benin OR Botswana OR Burkina Faso OR Burundi OR Cabo Verde OR Cameroon OR Central African Republic OR Chad OR Comoros OR Congo, Dem. Rep OR Congo, Rep. OR Cote d'Ivoire OR Equatorial Guinea OR Eritrea OR Eswatini OR Ethiopia OR Gabon OR Gambia OR Ghana OR Guinea OR Guinea-Bissau OR Kenya OR Lesotho OR Liberia OR Madagascar OR Malawi OR Mali OR Mauritania OR Mauritius OR Mozambique OR Namibia OR Niger OR Nigeria OR Rwanda OR Sao Tome and Principe OR Senegal OR Seychelles OR Sierra Leone OR Somalia OR South Africa OR South Sudan OR Sudan OR Tanzania OR Togo OR Uganda OR Zambia OR Zimbabwe ) |

**Table 2:**
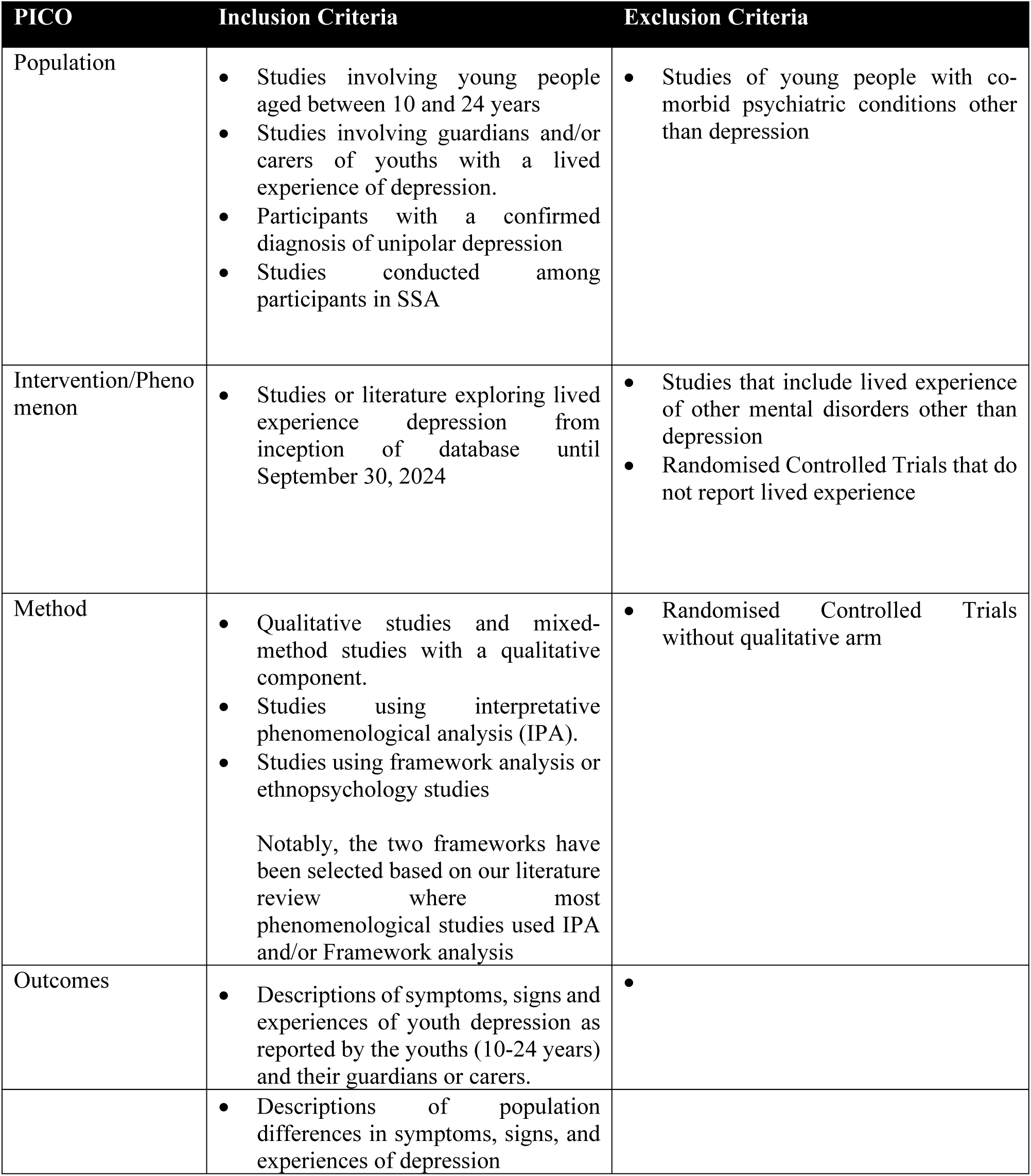
Eligibility Criteria.

| PICO | Inclusion Criteria | Exclusion Criteria |
| --- | --- | --- |
| Population | • Studies involving young people aged between 10 and 24 years<br>• Studies involving guardians and/or carers of youths with a lived experience of depression.<br>• Participants with a confirmed diagnosis of unipolar depression<br>• Studies conducted among participants in SSA | • Studies of young people with co-morbid psychiatric conditions other than depression |
| Intervention/Phenomenon | • Studies or literature exploring lived experience depression from inception of database until September 30, 2024 | • Studies that include lived experience of other mental disorders other than depression<br>• Randomised Controlled Trials that do not report lived experience |
| Method | • Qualitative studies and mixed-method studies with a qualitative component.<br>• Studies using interpretative phenomenological analysis (IPA).<br>• Studies using framework analysis or ethnopsychology studies<br>Notably, the two frameworks have been selected based on our literature review where most phenomenological studies used IPA and/or Framework analysis | • Randomised Controlled Trials without qualitative arm |
| Outcomes | • Descriptions of symptoms, signs and experiences of youth depression as reported by the youths (10-24 years) and their guardians or carers. | • |
|  | • Descriptions of population differences in symptoms, signs, and experiences of depression |  |

### Study selection

Using the PRISMA guidelines (15), the flow diagram in figure 1 illustrates the study selection process for this review. A research assistant (RA) ran the search strategy in the respective databases and identified 378 papers. The distribution of the identified papers based on their source were as follows: 18 from PubMed, 275 from Google Scholar, 18 from CNAHL,2 from Scopus and 65 from Embase. The RA checked all the 378 articles by reading through their titles. After removing 13 duplicates, we remained with 366 articles which were taken to the next level of screening where their abstracts were carefully read by the research assistant. This stage was done with supervision by PM. Out of the 366 articles, the RA removed 315 articles because they did not meet our inclusion criteria upon analyzing their respective abstracts. The reasons for removing these articles were explained to PM and thus, only 51 articles were taken to the next phase where PM and WM independently read full articles and assessed their eligibility. Disagreements between PM and WM regarding the suitability of a particular article were discussed between themselves and where a consensus was not reached, one of the supervisors, MU, was consulted for guidance. To that effect, only one article was disputed, and MU came in for a tiebreaker. After a full-text assessment of the 51 papers, 14 articles unanimously met the inclusion criteria and were thus included for qualitative synthesis and meta-aggregation.

### Assessment of methodological quality and Risk of Bias Assessment

Using the Critical Appraisal Skills Program checklist (CASP) for qualitative studies (16), PM and WM independently assessed the quality of the selected articles and the results for this exercise have been reported as part of our findings. Among other things, PM and WM also checked for any biases in the way the studies were done and reported.

### Data extraction, Management, and Synthesis

Under the supervision of MU, JJ and MK, the two researchers (PM and WM) independently extracted data from the selected articles. The extraction exercise was done in multiple phases whereby, a codebook for each paper was developed by each researcher (PM and WM). This codebook contained details like the study population, phenomenon of interest, the context, relevant methodological information and the study findings/results on one column and codes on another column. The coding process involved a line-by-line reading of the result sections of the selected papers. After independently coding each article separately, PM and WM had debriefing meetings to discuss and synchronize their individual codes to come up with a single codebook for each article/report. This ensured that there was reflexivity. A coding matrix for all the codebooks was then developed to allow for cross-comparison during data synthesis. This coding matrix included a code, its description and illustrations of the codes. Suffice to mention that during data extraction and analysis, our goal was to identify common themes, coping strategies, and the impact of socio-cultural factors on the lived experience of depression among adolescents/ youths and their guardians in SSA

Our systematic review and meta-aggregation used the following steps as proposed by the Joanna Briggs Institute (JBI) (17):

1. We gathered all findings (codes and themes) from all the reports reviewed, including accompanying illustrations.
2. We then categorized findings based on similarity in wording or concept with a minimum of two findings per category.
3. Finally, we developed one or more synthesized findings of a minimum of 2 categories.

Notably, we defined findings as all themes, metaphors and other analytic or observational data from the papers by the authors (17). Additionally, the category descriptions and the synthesized findings with their subsequent descriptions were created by a consensus process between PM and WM.

## Findings

### Study Characteristics

A total of 14 studies have been analysed and synthesized for this review. Table 3 shows the characteristics of the include studies. Notably, the studies were published between 2009 and 2024 and spans across 8 countries of Malawi, South Africa (4 studies) and Zimbabwe in Southern Africa, Kenya (3 studies), Uganda (2 studies), Rwanda and Tanzania in Eastern Africa and Nigeria in West Africa. The total sample size for all the reviewed studies includes 610 adolescents, 297 caregivers and parents, 1 partner, 70 stakeholders including mental health professionals, social workers, other healthcare providers and policy makers. For the adolescents or young people, there were 440 females,140 males and 30 whose sex was not recorded and their age ranged from 10 years to 24 years. Of the 297 caregivers, only 96 were male, representing 32%. Out of the 610 adolescents included in the sample, 219 were living with HIV, representing an HIV prevalence of 35.9% among study participants. Of the 14 studies, 6 were done among perinatal adolescents (18–23), 7 studies were done among young people living with HIV (5,19,24–28) and one study was done among internally displaced adolescents, living in camps and affected by civil war (29). Different theoretical models and qualitative study approaches were utilised among the selected studies including Grounded theory (5,6), Theoretical Framework of Behaviour Change (19), Ethnography (29), phenomenology (28), Consolidated Framework for Implementation Science (CFIR) (28), Syndemic Theory (20), Framework Analysis (23), Behavioural Model for Vulnerable Populations (21) and Interpersonal Psychotherapy Framework of Problem Areas (19).

**Table 3:** Characteristics of included Studies.

| Paper | Methodology/<br>Method/Analysis | Phenomena of<br>interest | Setting/Geographical/<br>Cultural | Participants | Author conclusions |
| --- | --- | --- | --- | --- | --- |
| Chukwuere et al (2022) | A qualitative, exploratory, descriptive design,<br>Purposive sampling, Semi-structured face-to-face individual interviews, analysis: Tesch's open-coding | Experiences of adolescents and their parent/guardians on the mental health management of depression | Participants recruited from two mental health care institutions and two mental healthcare units of two general hospitals in North West Province, **South Africa**. | • 32 participants (18 adolescents and 14 parents)<br>• Adolescents: 14 Females, 4 Males; 14-17 years old<br>• Parents: 13 female, 1 Male<br>• Clinical characteristic: adolescents had a diagnosis of depression | The devastating experiences of adolescents with depression and their guardian indicates a dire need for attention on their plights to facilitate adolescents' recovery and strengthen their coping skills |
| Judith Osok et al (2018) | An engagement interview approach, Grounded theory method was applied for qualitative data sifting and analysis<br>Convenient sampling, In-depth semi-structured interviews were done | Exploration of various practical, psychological, interpersonal, and cultural barriers to life adjustment, service access, obtaining resources, and psychosocial support related to pregnancy | • This was part of a larger prenatal depression project (which aims to study prevalence of depression and mechanisms for depression in pregnant adolescents)<br>• Participants were recruited from a rural Antenatal clinic of Kangemi primary health facility in Nairobi, **Kenya** | • 12 pregnant adolescents (ages 15–19)<br>• Clinical characteristic: positive PHQ-9 (cut off score was 15); 5 had severe depression, 7 had mild to moderate depression<br>• 3 caregivers and 1 partner who accompanied the adolescent for check-up to get a sense of the family's experiences and perspective | Findings have implications for service planning, including developing more integrated mental health services for pregnant adolescents. |
| Nicola Willis et al (2018) | In-depth interviews and Body mapping exercise were utilized. Participants created a painted map of their body to express their somatic and emotional experiences. Purposive sampling, semi-structured interviews | Explored the experience and manifestation of depression in adolescents living with HIV to inform intervention development. | • Participants were recruited from the Zvandiri ('As I am') programme-a model of differentiated clinical service delivery for HIV positive children and adolescents in **Zimbabwe** (www.africaid-zvandiri.org). | • 21 Adolescents living with HIV, diagnosed with major depressive disorder ( 15-19 years).<br>• Male -11, female 10<br>• 19 were orphans,<br>• All participants were diagnosed with Major Depressive Disorder | An understanding of HIV positive adolescents' own narratives around depression can inform the development and integration of appropriate mental health interventions within HIV care and treatment programmes. |
| Caroline Kuo et al (2020) | 8 FGDs with adolescents and their caregivers and 25 IDI with clinicians, researchers and other mental health providers,<br>Stakeholders sampled through snow bow sampling, sampling od adolescents and their guardians not documented<br>Thematic analysis: Open-coding, axial coding, and coding of marginal remarks | Identification of context-specific risk factors for family depression, and an exploration of family interactions around depression | Study done in Khayelitsha, one of the largest urban townships in **South Africa**<br>Recruitment from clinical sites serving HIV infected individuals and at a community-based organization working with youth at risk for HIV | 30 Adolescents, 13 to 15 years.<br>27 Caregivers of adolescents, 20-61 years<br>25 stakeholder, 18 years and above, 3 years working experience | This study will guide the development of ‘Our Family-Our Future’, a resilience-focused family intervention to prevent adolescent depression (ClinicalTrials.gov #NCT02432352). |
| Scholastic Ashaba (2019) | Semi-structured interviews: FGD and IDI utilised<br>Purposively sampling. Thematic analysis. Used grounded theory to develop a conceptual theory | An understanding of psychosocial challenges facing ALWH in rural Uganda and their effects on mental health and HIV treatment outcomes,<br>Findings were used to create a conceptual model linking the constructs that emerged, from the production of HIV stigma to its negative impacts on ALWH. | Recruitment was done in Mbarara Regional Referral Hospital (MRRH) HIV clinic, psychiatry ward, and the local community site, Nyakabare parish in **Uganda** to obtain diverse perspectives on mental health problems faced by ALWH and HIV-affected adolescents in the local community | FGDs: 30 Adolescents (13 to 17 years, no gender category): 6 were emancipated, 23 were living with HIV, 7 HIV status unknown<br>25 caregivers of ALWH: all female, 8 from MRHR HIV clinic (MHC), 9 from MRHR Psychiatric ward (MPW) and 8 from the community<br>IDI: 10 adolescents (2Male, 8 female) and 30 caregivers (5-MHC, 15-MPW, 10 community) | Interventions to correct community misperceptions about HIV can potentially reduce stigma and thereby improve physical and mental health outcomes of ALWH. |
| Obadia Yator et al (2022) | Theoretical framework of behaviour change was used.<br>purposive sampling, Focus group discussions and in-depth interviews were utilised. Thematic Analysis | The experiences and behavioural changes due Group Interpersonal Psychotherapy (IPT-G) delivered by CHWs among depressed postpartum adolescents (PPAs) living with HIV. | Participants recruited from two sites: Kangemi health centre and Kariobangi health centre in **Kenya** | 24 PPAs (FGD=19, IDI =5) aged 15–24 years old and living with HIV<br>CHWs (FGD= 7, IDI = 10)<br>Clinical characteristic: PPAs with a diagnosis of depression (scored > 10 on the Edinburgh postnatal depression scale) | Most of the PPAs reported an improvement in their interpersonal relationships with reduced distress and lessening of HIV-related stigma. |
| Theresa S. Betancourt et al (2009) | Ethnographic study.<br>Free listing interviews and Key Informant interviews (KIIs)<br>Purposive sampling | Exploration of local perceptions of psychosocial problems among children and adults to understand the problems affecting local children from their viewpoint and that of their caregivers | Participants from Echoli ethnic group, from Uganda, that has been displaced by the war in northern **Uganda** and were living in Awer and Unyama camps | Free listing interviews had 30 Adolescents: 14F, 16M, 10 to 17 years and 15 adults (12F, 3M)<br>KIIs had 25 Adolescents: 10 to 17 years (13F, 12M) and 32 adults (16F, 16M)<br>All the adolescents had lived in the camps for at least 3 months | The descriptions of these local syndromes are similar to western mood, anxiety and conduct disorders, but contain some culture-specific elements |
| Tasiana Njau et al (2024) | Phenomenological approach informed by CFIR Framework, Deductive thematic analysis | An exploration of implementation barriers and facilitators to formal mental health intervention-implementation among ALWHIV | Participants were drawn from HIV Care and Treatment Centers (HIV-CTCs) in Dar es Salaam, **Tanzania** | 15 adolescents (8 F, 7 M), 12-19 years<br>15 caregivers/parents, 27-61 years, (3M, 12F)<br>15 Healthcare providers (8 clinicians, 7 nurse-counsellors) | The findings of this study suggest an urgent need for an intervention that will be implemented to address depression specifically for ALWHIV, and that can be delivered in an integrated HIV care and treatment facilities in Dar es Salaam Tanzania and possibly similar settings in SSA |
| Zoe Duby et al (2012) | Using Syndemic theory, | An exploration of the subjective experiences of mental health and sexual and reproductive health among AGYW | Part of the HERStory study, evaluating a comprehensive intervention for AGYW in 10 districts in **South Africa**. Recruited from schools and communities | 237 AGYW aged 15 to 24 years | Considering the overlaps and interactions between mental health and SRH amongst AGYW, it is critical that mental health components are integrated into SRH interventions |
| Sally Field et al (2020) | Semi-structured interviews, Framework analysis used | Exploring the barriers and facilitators to service access for adolescents in Low-Resource setting | Adolescents from low-resource settings in Cape Town, **South Africa**. Prior to the study, participants had engaged with mental health services that were integrated into maternity care. | 12 female adolescents, 15 to 19 years old | Several key components for adolescent-friendly mental health services emerged from findings: |
| Lola Kola et al (2020) | Qualitative study, guided by the Behavioural Model for Vulnerable Populations<br>Purposive sampling used.<br>Content analysis | An exploration of how predisposing, enabling, and need factors impact the health services utilisation experiences of perinatal adolescents with depression | Participants were respondents of a Randomized Controlled Trial (RCT) conducted in 28 primary health facility in 11 local government area in Ibadan, **Nigeria** | 17 females (they were depressed, pregnant, adolescents at the time of the RCT); 21 to 23 years old<br>25 caregivers: 22F, 3M 44 to 54 years old | Participants identified unsupportive and stigmatizing clinic environments towards pregnant and parenting adolescents as significant barriers to accessing available care. |
| Manasi Kumar et al (2022) | Interpersonal Psychotherapy Framework of problem areas guided the the semi-structured interviews<br>Analysis done using two-stage process-manual coding and then through triangulation via NVivo | An understanding of Interpersonal challenges facing peripartum adolescents experiencing unplanned pregnancy and depression | Participant recruitment happened in two Primary care clinics in Nairobi, **Kenya**. | • 23 Peripartum adolescents; 16 to 18 years<br>• Clinical characteristics: 70% screened positive on Edinburgh Postnatal Depressive Scale (EPDS): 13% severely depressed, 39% were moderate to severely depressed | Our interviews deepen understanding of peripartum adolescent mental health focusing on four IPT problem areas. The Interpersonal Framework yields meaningful information about adolescent depression and could help in identifying strategies for addressing their distress |
| Theresa T Betancourt et al (2011) | Qualitative methodological approaches: Free listing (FL) interviews, Key Informant interviews (KIIs) and Clinician Interviews<sup>1</sup> (CI)<br>FL participants selected based on the principle of “maximum variation” KIIs sampled using snowball. | Exploration of common mental health problems and their symptoms/indicators among HIV-affected youth in Rwanda | Most participant selection took place in the waiting area of the Rwinkwavu District Hospital’s Infectious Disease Clinic. | • FL participants: 43 adolescents (10 to 17 years, 20 Females, 23 males) and 31 adults (13 females, 18 males)<br>• KIIs in 2007: 36 adults (11 females, 25 males) and 38 adolescents (13 female, 25 males)<br>• KIIs in 2009: 44 adults (18 females, 26 males) only<br>• CI Participants: 10 adults (6 females, 4 males) | Our findings underscore the importance of understanding local manifestations of mental health syndromes when conducting mental health assessments and when planning interventions for HIV/AIDS-affected children and adolescents in diverse settings. |
| Bradley N. Gaynes et al (2024) | Semi structured interviews, purposive sapling | Exploration of counselling and peer support delivery preferences amongst adolescents living with HIV (ALWH) and Experiencing Depression | This is part of the HIV Engagement and Adolescent Depression Support (HEADS-UP) study in Malawi | • 25 Adolescents living with HIV; 12F, 13M, 13- 19 years old, diagnosed with depression (Beck Depression Score $\geq 13$,<br>• 8 Stakeholders<sup>2</sup> $\geq 18$ years old=<br>• 10 Periscope Participants & Implementors<sup>3</sup> 18-24 years | This pilot study will provide insights into youth-friendly adaptations of the Friendship Bench model for ALWH in Malawi and the value of adding group peer support for HIV care engagement |

F= Female, M = Male
<sup>1</sup> Mental Health Professionals, pediatricians and social workers
<sup>2</sup> Caregivers, Clinicians, Ministry of Health Officials, Clinical Staff
<sup>3</sup> Previously Participated in, or implemented, a Friendship Bench Intervention for Perinatal Women Living with HIV in Malawi

### Methodological Quality

Table 4 shows a summary of the quality appraisal rating, using the Critical Appraisal Skills Program (CASP) tool (16). From the table, the quality ranged from 80 to 100% on the appraisal tool. All the studies had a clear statement of the study aims. However, only one study explicitly mentioned that they were exploring the experiences and manifestation of depression in adolescents living with HIV (26). Otherwise, most of the studies’ objectives and titles did not explicitly mention of the lived experience of depression among the youth. The studies explored either adolescents’ experience regarding the barriers they face in accessing care for depression (6,19,21,23,28), contextual issues around youth depression and family interactions around adolescent and maternal depression (27), adolescent’s psychosocial issues in the context of HIV, Depression and adolescent or teen pregnancy (19,24), and experiences regarding management or interventions that addresses youth depression (24,30).

**Table 4:** Quality Appraisal Ratings using CASP.

| Paper | Q1 | Q2 | Q3 | Q4 | Q5 | Q6 | Q7 | Q8 | Q9 | Q10 | Total (/10) |
| --- | --- | --- | --- | --- | --- | --- | --- | --- | --- | --- | --- |
| Chukwuere et al (2022) | Y | Y | Y | Y | Y | Y | Y | Y | Y | Y | 10 |
| Judith Osok et al (2018) | Y | Y | Y | Y | Y | Y | Y | Y | Y | Y | 10 |
| Nicola Willis et al (2018) | Y | Y | Y | Y | Y | Y | Y | Y | Y | Y | 10 |
| Caroline Kuo et al (2020) | Y | Y | Y | CT | Y | CT | Y | Y | Y | Y | 8 |
| Scholastic Ashaba (2019) | Y | Y | Y | Y | Y | CT | Y | Y | Y | Y | 9 |
| Obadia Yator et al (2022) | Y | Y | Y | Y | Y | Y | Y | Y | Y | Y | 10 |
| Theresa S. Betancourt et al (2009) | Y | Y | Y | Y | Y | Y | Y | Y | Y | Y | 10 |
| Tasiana Njau et al (2024) | Y | Y | Y | Y | Y | Y | Y | Y | Y | Y | 10 |
| Zoe Duby et al (2012) | Y | Y | Y | Y | Y | CT | Y | Y | Y | Y | 9 |
| Sally Field et al (2020) | Y | Y | Y | Y | Y | CT | Y | Y | Y | Y | 9 |
| Lola Kola et al (2020) | Y | Y | Y | Y | Y | N | Y | Y | Y | Y | 9 |
| Manasi Kumar et al (2022) | Y | Y | Y | Y | Y | N | Y | Y | Y | Y | 9 |
| Theresa S Betancourt et al (2011) | Y | Y | Y | Y | Y | N | Y | N | Y | Y | 8 |
| Bradley N. Gaynes et al (2024) | Y | Y | Y | Y | Y | CT | Y | Y | N | Y | 8 |
| % Yes | 100 | 100 | 100 | 93 | 100 | 43 | 100 | 93 | 93 | 100 |  |

All the studies were qualitative in nature, thus semi-structured in-depth interviews (IDI) and focus group discussions (FGDs) were used. Only two studies (25,29) used Free Listing techniques in addition to IDI and FGDs. The recruitment strategy in all the studies was clearly stated and appropriate. However, one of the most challenging areas in most of the studies was that the authors did not clearly state any potential bias and their influence, if any, during research question-formulation and data collection. Furthermore, except in one study, there was no mention of any event that may have happened during the study. Ethical principles were considered in all the studies and it worth noting as well, that the data analysis in all the studies was sufficiently rigorous. Authors included at least one participant’s quotes per theme to illustrate their developed themes. However, in two studies, where free listing was utilised, instead of quotes, the authors provided a list of local terms that were mentioned as part of the symptoms that form a cluster of symptoms of locally, culturally and contextually relevant syndromes of distress (25,29).

### Synthesised Findings On the Lived Experiences Of Depression By The Youth

Related findings from this review were grouped into eleven different categories. The related categories were grouped to form three main synthesised findings of Making sense of depression, Mental Health Systems and Services and Contextual issues/factors. Table 5 below show the synthesised findings and the categories for each synthesized finding.

**Table 5:** Summary of Meta-Aggregation Findings.

| SYNTHESIZED FINDINGS | CATEGORIES |
| --- | --- |
| Making Sense of Depression | 1. Lost in Space- State of Confusion<br>2. Communicating their feelings-Idioms of distress<br>3. Symptoms of Depression<br>4. Causes and Risk Factors for Youth Depression<br>5. Coping and Support |
| Mental Health System and Services | 1. Available Services and Interventions<br>2. Experience with Utilisation of Services<br>3. Barriers and Facilitators for Accessing Services for Depression |
| Contextual Issues | 1. Pregnancy-Adolescent Perinatal Depression<br>2. HIV-Adolescents Living with HIV and Depression<br>3. Sociocultural Issues |

### Synthesized finding 1: Making Sense of Depression

The fist synthesized finding of this meta-aggregation explores an understanding of what the youth in SSA think about depression. It looks at the meaning they attach to depression or their interpretation of being depressed, the symptoms they experience, what they think are the causes of depression as well as the coping mechanism and support they get from their significant others and their immediate environment.

#### Lost in Space-Going through a confusing phase of Life

The first category under this synthesised finding is ‘lost in space’. It was found that most of the youth, especially those who were experiencing depression for the first time were taken unawares of what was happening with them and they looked ‘lost’ and not interested in life as observed by an adolescent from Cape Town, South Africa (23): *“.…they just look lost. Like they living life not really wanting to live”* (18-year-old, female Adolescent, South Africa) *(23)*.

Some are confused and pessimistic about the future, given their state of mental health: *“ªFor now there is nothing [in the future]. It’s just hazy and a bit complicated to understand where it’s headed to. At times it’s just sorrowful and just so sad”* (18-years-old male adolescent, Zimbabwe) (26). The youths are even more confused because sometimes, when reality hits them, they are aware that they are not feeling well. However, it becomes difficult for them to figure out whether what they are experiencing is just a normal period of adolescence or there is more to it. They personalize their experience, and this makes things even worse. Their mental health keeps dwindling till it impacts on their personality: *‘It is worrying me a lot because I am a fun person. I love laughing a lot and making the people around [me] very happy, but all those things changed because I cannot cope with this condition, and it is very discouraging. It is really so strange to me because most times, I feel powerless, I cannot handle.’* (Adolescent 2, NW-South Africa) (30)

#### Communicating their feelings-Idioms of Depression

The next category in this synthetic finding was to do with how the youth communicate their experiences to their peers, family and their significant others. We observed that across the sub-Saharan African region, there is a linguistic complexity when it comes to communicating and describing most psychiatric disorders including depression and this is influenced by culture, which varies across and even within the same country (25,27,31). The linguistic complexities are obvious with use of idioms of distress or depression. In many countries in SSA, depression is often used interchangeably with the word ‘stress’. *“I only know that when someone is stressed it is called depression”* (Adolescent, South Africa) and *“People who are depressed, they call it stress”* (Parent, South Africa) (27). This was confirmed by a social worker: *“There’s no word in Xhosa that describes depression. It’s depression in English, but in Xhosa there’s no word that people associate with [depression], I don’t know, that explains the depression. People use different words”* (Social Worker, South Africa). Other idioms of distress/depression in other countries include being entrapped in a small world: *They become so sad and hate everything around them. The whole world becomes like a box [feel trapped]. Some children may commit suicide by hanging or poisoning themselves”*(31). ‘Thinking deeply’ (locally known as *kufungisisa* in Zimbabwe), pain and darkness (26). In some countries like Uganda and Rwanda, within the indigenous system, depression does not exist in its entirety as it is known in the western world as defined by the ICD and DSM systems. In both Uganda and Rwanda, there are local syndromes which look like depression but have cultural-specific features that define and distinguish them from the DSM’s mood disorders like depression and anxiety (25,29). Briefly, in Rwanda, they have three syndromes of *guhangayika*, *agahinda kenshi* and *kwiheba* which are equivalent forms of child and adolescent depression with some features of anxiety (25). In Uganda, *two tam* (depressive and anxiety features)*, kumu (*often associated with features of depression, marked by severe loneliness and siting while holding both cheeks in the palm*) par* (depressive symptoms with prominent externalising/anti-social behaviours) (29). These syndromes have similarities in almost all the symptoms, what distinguishes them from amongst each other is the severity of the symptoms within a particular syndrome. For example, in Northern Uganda, *two tam* share similar features with *kumu*, only that *two tam* has more severe features with presence of suicidal behaviours (29). In Rwanda, *Guhangayika* is a mild form, while *agahinda kenshi* is the moderate form and *kwiheba* is the severe form of the equivalent of depression (25).

Apart from communicating their feelings and mood through talking, some youths use arts, specifically, drawings, to express their feeling to their significant others with the hope that they will be heard. For example, one 16-year-old boy from Zimbabwe drew a chicken, when asked to draw his experience of depression. He drew a hen, to represent his current state of emotion: *“Because anytime it [hen] can be killed. So just like me, l am like a hen. l don’t know when l will die but I just know that l will die because of the situation that l am in”* (Male Adolescent, Zimbabwe). Apart from arts, others used writings as a medium of communication. *‘I feel my mother understands me now. I definitely feel so because she told me that she read what I have written and realised that she was not listening to me enough*’ (Male Adolescent, South Africa).

#### Symptoms of Depression

Making sense of depression also includes the common symptoms or features of depression that are experienced by the youth in SSA. We include common symptoms that appeared in at least two reviewed papers. These symptoms fit into the three main domains of cognition (thoughts), feelings and behaviours (actions). The most common symptom was mentioned in 8 reviewed papers, and this was sadness or sorrow (18–20,22,23,25,26,31). The second most common symptoms were mentioned in 7 studies and these included social withdrawal or isolation (18,19,23,25,27,30,31) and hopelessness (19–22,25,30,31). The other common symptoms mentioned in at least two reviewed papers included excessive crying (18,23,25,26,28,30), suicidal thoughts and behaviours (18,19,22,30,31), overthinking (19,25,28,30), non-somatic pain (18,27), loss of appetite (18–20), anger or irritability (19,25,31), changes in sleep pattern (18,28,31), negative thoughts (19,28), low self-esteem (22,31), stress (18,30) and somatic symptoms like headache, stomach-ache and chest pains (18,28). Table 6 below shows an index of the symptoms that were mentioned in the reviewed articles and table 7 shows some quotes illustrating some symptoms as expressed by the youths themselves.

**Table 6:** Common Reported Symptoms of Youth Depression.

| Symptom | Frequency: Number Of papers appearing |
| --- | --- |
| Sadness or sorrow or low mood | 8 |
| Social Withdrawal and Isolation | 7 |
| Hopelessness | 7 |
| Anger | 3 |
| Pain (non-somatic) | 2 |
| Stress | 3 |
| Overthinking or thinking too much | 4 |
| Excessive Crying | 6 |
| Appetite Changes | 3 |
| Suicidal thoughts or Suicidal attempt | 5 |
| Changes in Sleep Pattern | 3 |
| Somatic Symptoms (Headache, Stomach-ache Chest Pain) | 2 |
| Negative Thoughts | 2 |
| Low self-esteem | 2 |

**Table 7:** Descriptive Quotes illustrating some symptoms.

| Symptoms | Descriptive quotations |
| --- | --- |
| Hopelessness, suicidal behaviours | *'I tried committing suicide because I feel hopeless, I was losing my mind and I feel killing myself will bring to an end all the suffering in this world,...is not interesting at all.'* (Female Adolescent, South Africa) (30) |
| Sadness, Anger | *"I get very sad...I'm trying...I'm telling them that...and they say...you're weak. It makes me angry"* (female Adolescent, South Africa)(20) |
| Changes in sleep pattern, loss of appetite and social withdrawal | *"Right now, she sleeps a lot and does not eat, she is always indoors"* (a guardian of a depressed, pregnant adolescent; Kenya) (18) |
| Social withdrawal, Stress | *"I used to sit in the house and had no friends. But when I started coming to this [therapy] group, I found friends here, outside I have also made friends. I used to be stressed"* (Female youth, Kenya) (19) |
| Isolation/withdrawal | *"I was afraid of everything and sometimes ran away because I wanted to be alone. My aunt suggested I see a doctor for crazy people. I thought, dah! So, I am crazy! I have HIV; and on the other hand, I am crazy ... I did not go. You* |
|  | *know it is only crazy people who go there (mental health care unit), which means I will be one of them.” (28)* |
| Non-Somatic (Psychological) Pain | *“They first start with going to the clinic. In the clinic they present with headaches and whata, whata, whata and the doctor will say, “Nothing’s wrong with you.” Gives you pain blocks. The client will then say, “But I still feel this pain despite this pain block.” (Psychologist, South Africa) (27)* |

#### Causes or risk factors of Youth Depression

This review found various issues that the adolescents, their guardians, as well as stakeholders identified as risk factors and/or causes of youth depression in the SSA region. The most identified causes of depression included poverty, chronic physical health conditions like HIV, domestic and sexual violence, pregnancy and early motherhood, disrupted family system, HIV in the family, parental ill-health especially maternal depression, adverse childhood, poor interpersonal relationships with peers and family members, orphanhood and being a surrogate child, socio-political instability (25,27,30). In some tribes, like in South Africa, some people attributed depression to witchcraft: “*Many people believe a person with a mental problem has been bewitched, so it is not even easy to say if that is depression because once you see persistent negative thoughts and behaviours, you cannot explain, plus the fact that our expertise to diagnose it is low, we end up losing them to witchdoctors”*(28).

#### Coping Strategies and Support

Before reaching out for support from others, the youth try different ways to cope with the depression. Some of the coping strategies that our review found include writing, being on the phone and solitude (20,21,27,30); “*When I come back from school, I love staying in my room alone maybe writing down how I am feeling or with my phone. I do not like talking to anyone*.’ (Male Adolescent South Africa) (30). Suffice to mention that solitude was the most common coping mechanism adopted by most youths from different countries and was mentioned in several reviewed papers (20,21,27,30). Ignoring people’s comments and attitude towards their depressive episode and staying focused on getting better, was also used as a coping method especially in situations where they were being stigmatised (21): “*I continued going to see matron and didn’t pay too much attention to discouraging comments from people”* (adolescent mother, Nigeria) (21). Religion is another way through which adolescents with depression cope (31). It was found that through religious gatherings, young people with depression get some social connectedness, through religious sermon, like in Christianity, young people’s hopelessness is restored by among other things, drawing lessons and motivation from bible characters who endured challenging moments (26,31). In the context of young people living with HIV (YPLWH), positive role-modelling was also noted as a coping mechanism as it is used to instil hope among YPLWH (31).

While utilising the above-mentioned self-help strategies, the young people reach a phase where they feel like they have had enough as mentioned by a female adolescent: *“I cannot cope with this condition, and it is very discouraging. “It is really so strange to me because most times, I feel powerless, I cannot handle”* (Female adolescent, South Africa). In special contexts like in adolescent perinatal depression or depression in adolescents living with HIV as it will be discussed further, coping is even more challenging as in those contexts, there is a double burden of stigma. “*They say having a child is a good thing, but as a teenager it is a burden, it’s difficult to cope”* (Pregnant Adolescent, South Africa) (20).

While trying to cope using the different methods outlined above, the youth, eventually, reach a point of no return and are willing to be supported (27,30). The support is usually in form of learning new ways to cope and adapt, having a platform to vent and release their emotions or even to get social support (24,27,30). The support usually comes from peers, family members, religious institutions and many more (24,27,31). The probable attribution of the depression was noted to be a determinant of getting support from family. For example, when adolescents are depressed due to pregnancy-related stressors, they rarely get support from the family and oftentimes, they are ostracized and disowned by the family (19,21,22). While some family members are reportedly unsupportive, others are supportive and empathetic towards the youth (18): *“I have realized that I am her only friend, and I do not want her to be alone. We are all okay with this as parents and my mother-in-law supports me”* (Mother of a depressed pregnant Adolescent, Kenya). Sometimes lack of support emanates from lack of knowledge about mental health and depression, in particular. *‘No one understands me; they are not concerned about my emotional health. My dad is not listening to me, he does not care how I feel, it makes things difficult for me, I feel lonely, I have tried killing myself many times. My home is not favouring my emotional health; it hurts me a lot”* (Female adolescent, South Africa) (30)

### Synthesized Finding 2: Mental Health Systems and Services

The second synthesized finding looks at the health system, with particular emphasis on the nature of the available services, the experience of the youths while utilizing such services and finally, the barriers that the youth face and the opportunities at their disposal in accessing such services.

#### Available services and Interventions

Our review found that different services exist in SSA for the youth with depression and their guardians. The services range from screening and diagnostic services to psychosocial services including counselling services. These services in some instances are underutilised because of barriers to be discussed later. In SSA, screening and diagnosis for depression is not routinely done unless it indicated (28). But then, even when indicated, it is rarely done because of lack of expertise among primary healthcare workers: “*The biggest challenge is the lack of intervention skills and expertise to diagnose depression. Our clinic does not have a psychologist to ensure they [adolescents] get treatment* (Nurse counsellor, Tanzania) (28). Additionally, validated screening tools are not always available for service providers to use*: “But there are not even appropriate psychological intervention guidelines or a simple [screening] form that an adolescent can fill out that we can at least learn to use.”* (Clinician, Tanzania). Despite these challenges, when the screening is properly done, young people do appreciate its relevance, and the acceptability is good because most of them do not realise that they have depression up until they have been screened: *‘They [screening questions] helped me a lot … I think it’s to make sure, for you to be healthy, because also stress can affect your baby’* (18-year-old postnatal adolescent, South Africa) (23). However, there are mixed reactions from the youth upon learning of their depressive status following the screenings. While some are comforted by the fact that their phase of confusion or ‘lost in space’ is over as they are then aware of what was wrong with them, others become emotional about it as mentioned by a 19-year-old postnatal adolescent: *‘… It was a very emotional process. It was when [after the screening] I realized I wasn’t happy with my pregnancy and I wasn’t happy with a lot of things, so it was sad.’* (Postnatal adolescent, South Africa) (23). The acceptability of the screening process depends on the approach of the personnel doing the screening: *“She [the screener] made you feel comfortable. It was like, I could tell her anything. So, the answers I gave her was just honest answers … It was quite simple. It wasn’t like that in too deep. Like she didn’t pry too much into your personal stories. She kept it on a normal level.’* (18-year-old pregnant adolescent, South Africa) (23).

Various psychosocial interventions are available in SSA. Our review identified three common psychosocial interventions which help youths with depression. These were counselling, support with education for school-going young people and vocational skills training for out-of-school youths (23,26). Counselling being the commonest, it is received with mixed reactions as well. Some youths are concerned with their privacy, while others are afraid of being judged: *‘I thought they were going to tell me about how young I am. I thought they judge a person or something, that was why I did not want to talk when I saw her [the counsellor] the first time.’* (postnatal adolescent-15, South Africa) (23). Because of this negative attitude against counselling, some develop resistance: “*For me it was like whatever, I’m just gonna go, yeah, whatever they tell me I’m just gonna say ‘mmm yea’ to everything, I’m gonna talk like that. And then when I went to see her [the counsellor] it was different.”* (postnatal adolescent-17, South Africa) (23). Perceived benefits of counselling included being listened to, having an avenue to release emotions by freely talking about their issues, being understood, feeling of being validated (23,26). Through their lived experiences, the youth have preference regarding the nature of a counsellor they would want to be attended to. Some of characteristic of their preferred counsellor include; one who is calm, warm, caring, a good listener, empathetic, welcoming, respectful, knowledgeable, not too formal, non-judgemental and one who upholds privacy and confidentiality (23,24): *“They should be calm, and also one who can keep secret of what they are being told; they shouldn’t tell it to another person”* (14-year-old Male, Malawi). Overall, the youth prefer mental health services that are youth-friendly, preferably, provided by peers or a professional who is not too old to understand youthfulness (24): *“Do not be using elderly people with the youth. It should be all about the youths so that the youths should be open…He or she shouldn’t be an elder above 40 going upwards, youths will be failing to be open”* (Friendship Bench Provider/interventionist, Malawi) (24). Furthermore, it was found however, that most of the youth, especially those living with HIV, prefer peer-led interventions (24,26). Some interventions were marred by linguistic complexities due to lack of cultural adaptations (27). Unlike having an all-size-fit-all kind of approach, it was noted that the youths preferred context-specific interventions, for instance, ALWH required special consideration regarding the content of the psychological interventions given to them (24). They proposed that interventions of depression for ALWH should include sessions on dealing with community misperceptions (31), disclosure issues, positive living and coping skills, among others (24). The acceptability of the interventions depends on the perceived benefit of the intervention (21).

#### Experience with utilization of services

The youths had a mixed feeling and experience with the healthcare system. Most of the participants in all the reviewed studies were not satisfied with the quality of services rendered to them and often, complained about the hospital environments and the healthcare providers as being unsupportive: *“Like the hospital should be friendly, it must help us to heal. I mean this environment is not friendly to us like the environment is not friendly like is just more depressing so I feel they should introduce things that would make us feel better.”* (female Adolescent, South Africa) (30). *“To me, I think the hospital should help us by being friendly to us like the nurses and doctors; they should try to understand what we are passing through emotionally. At least, when we come with our problem, we should be smiling while going home.’* (Male Adolescent, South Africa) (30). There were more complaints against some healthcare workers (HCW) as being harsh and inconsiderate as lamented by a guardian: *‘I do not just understand how they operated because I think they were against her [my daughter] like they were not treating her fine; they were disrespectful, because I used to tell them what was happening but they would not listen to me. It makes me feel so bad and sometimes, I will cry and eeeem what made me to cry was they were not listening to what I was telling them about my child[‘s] situation.’* (Female Guardian South Africa) (30). It was observed that some HCW lack listening skills, were not flexible and accommodative to the needs of the youths. For example, some HCW prioritise medication against the young people’s preference for psychotherapy: *“Doctors do not understand the psychological burden you have. They think medication is the most important thing. We often have different priorities; you (as an adolescent) think of the emotional issues, but doctors think of medication”.* (Adolescent, 15-year-old, Tanzania)*(28).* Furthermore, the youths complained of poor communicating skills by some HCW. The HCW however, complained that some youths with depression are uncooperative and difficult to handle: “*Those [depressed] young girls are very rude and difficult to at tend to. They are very disrespectful”* (HCW, Nigeria) (21). This tension between some HCW and depressed youths contributes to lost to follow up and in some instances, the youths tend to have favourite HCW to an extent that they only visit the hospital/clinic when their favourite HCW is available as mentioned by a perinatal adolescent: *“I went to the clinic only when I am sure [the] matron was on duty”* (Perinatal adolescent, Nigeria) (21). Even though some of these issues will be discussed under barriers to accessing healthcare service, one point worth mentioning on the experience of the youths in the utilization of mental health services is to do with how the mental health clinics are structured in some countries. It was observed that most health care services are scattered within the hospital so in situation where for example, an adolescent requires antenatal services and mental health services, or requires HIV care and mental health services, then they must move from one section of the hospital to the next (23,28). Sometimes they even must visit the hospital several times in a month to get other complementary services because of lack of a multidisciplinary clinic as most services are disintegrated within the hospitals (23,28).

#### Barriers and Facilitators for accessing Mental Health Services

##### (i) Barriers

Using the Consolidated Framework for Implementation Research (CFIR) theoretical framework (32), we identified and categorised the barriers into five domains as follows:

**Individual factors**-these include negative attitude of some HCW about youth mental health as well as the negative attitude of some young people themselves. “*Some of the nurses are so rude neh, and some of them like I remember my first time of going to [name of the hospital] neh, so I saw this social worker neh, she was so rude to me telling me how like she was saying I am a mama’s baby instead of motivating me.”* (Adolescent, South Africa) (30). Stigma by some HCW (21), lack of awareness about mental health by both the young people and their guardians, inadequate trainings and skills in detecting depression by some HCW(27,28): *“They could not figure out her problem at the clinic until doctors visited our school for mental health education on World Mental Health Day. I finally realised it was depression.”* (Guardian, Tanzania) (28) and *“We do not deal with mental health problems. The biggest challenge is the lack of intervention skills and expertise to diagnose depression”* (HCW, Tanzania) (28).

**Inner setting:** limited time to listen to the youths and attend to their needs, unstructured clinics, and non-therapeutic hospital environments (28).

**Outer setting:** lack of political will in addressing youth mental health by the governments. “*We have manuals for TB, for example. So why not for depression while they [government] know how serious this problem [depression] is*” (HCW, Tanzania) (28).

**Process factors:** long pathway to receiving formal care as some initially go for traditional or faith-healing before seeking formal healthcare from hospitals (27,28). Some use informal care due to the unavailability of formal services as mentioned by a guardian and a healthcare worker; “*I have exhausted all options. I started taking my daughter to traditional healers, pastors, and local counsellors on the street, but nothing seemed to help. She will be okay for one day and returns to her world the next day. If there is a possibility of getting the service here, it will be helpful”.* (Caregiver, Tanzania); and *“our expertise to diagnose it [depression], is low; we end up losing them to witchdoctors.”* (HCW, Tanzania) (28).

**Intervention Characteristics**: high cost of the interventions in some countries (30), lack of culturally sensitive tools and interventions (25,28). It was observed that most of the tools being used are simply translated versions of western-developed tools with some adaptations (25).

##### (ii) Facilitators

Three facilitators for utilization of formal mental health system were identified. Bilingualism among some youths was an enabler for seeking care as it was easier for such youths to understand both the indigenous and foreign concepts of depression (27). Perceived benefits of an intervention like counselling also played a role in facilitating the youth to seek and continue with formal care (21). Another facilitator is the increased in the use of World Mental Health Day to promote mental health awareness among the youths especially when such events are done in schools (28).

### Synthesized Finding 3: Contextual Factors/ Issues

The context in which a young person suffers from depression shape and determine how they experience the depression. We identified three main contexts of interest to be discussed and these include (i)Pregnancy-adolescent perinatal depression, (ii)HIV-Adolescent living with HIV and Depression (ALWH-D) and (iii)socio-cultural contexts.

#### Pregnancy- adolescent perinatal depression

Teen pregnancies are characterised by denial of the pregnancy by the partner, social stigma, financial burden to the teen as well as her family, ostracization, early motherly responsibilities, concerns with welfare and future of the unborn baby, regret, guilty, lack of support from the family due to stigma, community beliefs about teen pregnancy: *“Many of these girls are irresponsible and promiscuous and do not listen to parents…it is no wonder that they receive very little support from relatives. they need to learn the hard way”* (Care provider, Nigeria), (21) and “*an aunt told me it [teen pregnancy] is a sin.”* (15-year-old, 9 months pregnant, Kenya) (18), inability to distinguish between pregnancy-related morning sickness and morning symptoms of depression, lack of interpersonal relationships and intimate partner violence (18,20–23). All these factors interrelate with each other to predispose the adolescent to depression and perpetuate the depression. Eventually, it influences the health-seeking behavior of and service utilization by the depressed pregnant and postnatal adolescents. Support becomes an important issue because on one hand, the adolescents need to attend to antenatal care, on the other hand, they also need to get mental health support. Unfortunately, the support is minimal due to issues around stigma and worse still, the family’s reaction sometimes promotes suicidal ideation: *“I don’t know whether I am depressed the way you describe it, but I regret a lot ... (crying again). I went to a bridge at home and wanted to throw myself because my grandfather is very harsh and upset with me”* (pregnant adolescent, Kenya) (18).

#### Adolescent living with HIV and Depression (ALWH-D)

As is the case with pregnant adolescents, adolescents living with HIV (ALWH) and depression face numerous unique challenges due to the double stigma of HIV and mental ill-health. Through a qualitative study on Community beliefs, HIV stigma, and depression, among adolescents living with HIV in rural Uganda, Ashaba Scolastica et al (2019), developed a conceptual framework that links HIV and Depression in ALWH (31). According to the model, negative community perceptions (about aggression and presumed early mortality of ALWH) and vulnerability factors due to the HIV (like orphanhood, poverty, physical health challenges) predisposes ALWH to internalization of stigma, which eventually leads to depression and subsequently ART non-adherence, as illustrated in figure 2 below (31). The Non-adherence to ART is used by some ALWH as a means of suicide, termed as “Slow Suicide” by Nicola Willis et al (2018) (26). The normal developmental stage of adolescence is characterised by being concerned with the body structure and identity and in the general adolescent population, these predisposes them to depression. ALWH are even more concerned with their physical outlook because of the direct and indirect impact of the HIV: “*The pain that I go through each day I go to school because of my body…It’s small so people always tease me”* (ALWH,18 years old, Zimbabwe) (26). Orphanhood, because of HIV puts ALWH at an increased risk of developing depression (26,27). Orphanhood leads to ostracization which result in ALWH to keep moving from one relation to another thereby affecting their academic performance, thus, some of the ALWH miss school (26,27). The poor academic performance is not only due to ostracization, but also due to the direct impact of the HIV like multiple hospitalization, bullying and teasing in school. The result is school dropout and this in turns leads to a vicious cycle of HIV, poverty and depression.

**Fig 2:**
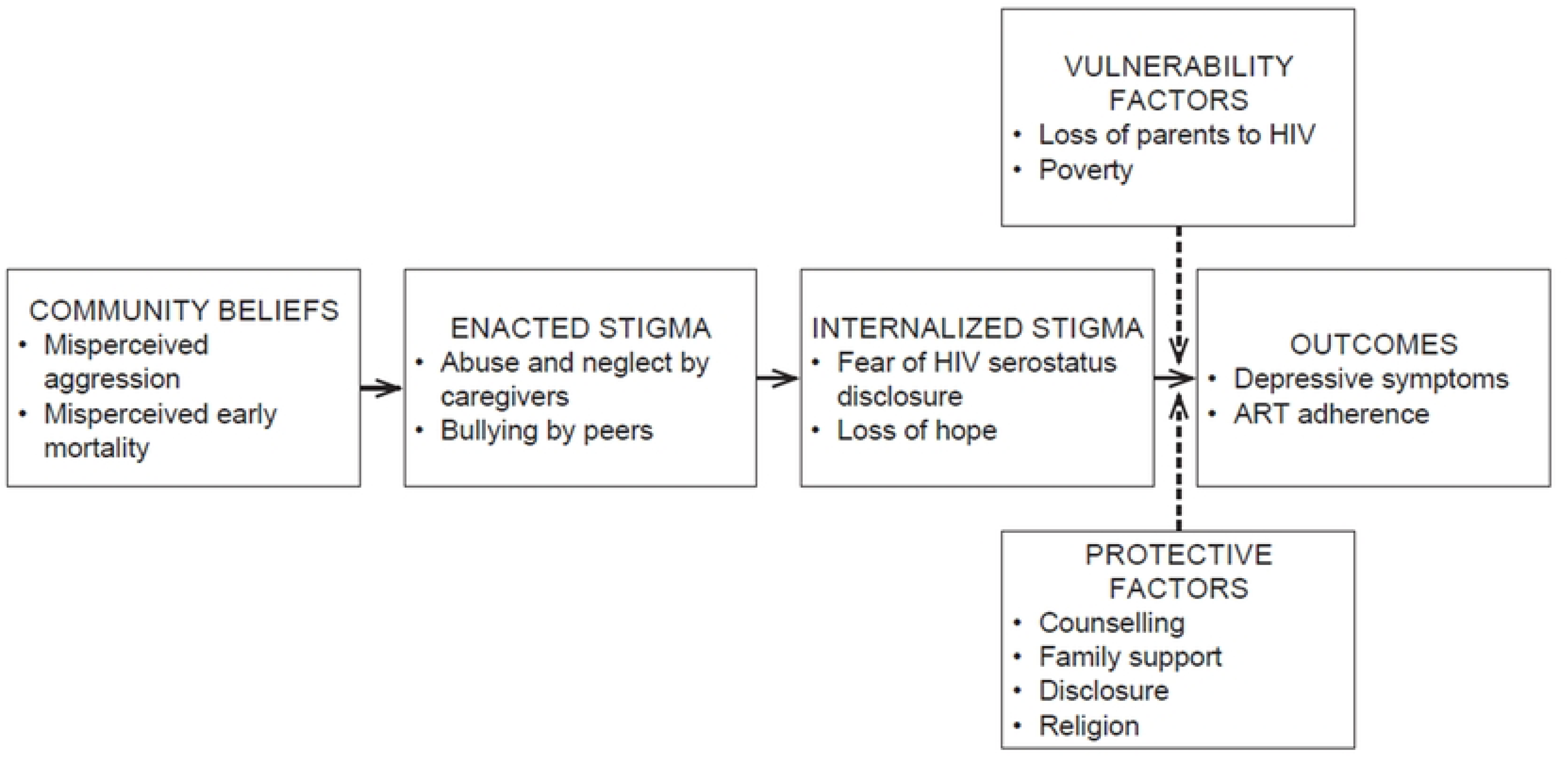
Conceptual model by Ashaba Scolastica et al (2019), linking community beliefs, HIV stigma, and health outcomes among adolescents living with HIV(31).

Another critical issue affecting ALWH is that of disclosure of their HIV status to their peers and the extended family(31). This collides with disclosure of their mental health straggle thus, they tend to internalise most of their straggles, as such, they indulge in high risky behaviours like unprotected sex, abusing alcohol and substances, self-harm and even committing suicide. Some of them see death as imminent because of the HIV so they would rather kill themselves than to wait for the natural process (26).

Due to poverty, some youths, especially, girls, also indulge into commercial sex to earn a living and in so doing, they end up being impregnated*. “I got pregnant in the course of hustling because at home I could not get what girls need and we were sleeping hungry all the time. I am the eldest and so if I bring [money] even my sisters will be alright. My father works but he is a drunkard and my mother washes clothes for more affluent people but it’s not enough…”* (17-year-old, pregnant adolescent, Kenya) (18). The pregnancy and the HIV status further predisposes them to depression or even perpetuates an already existing diagnosis of depression. The comorbidity of HIV and Depression, in the context of pregnancy creates what we would call, “the Triple Burden of Stigma” which signifies increased stigma leading to increased distress. HIV in the family was also noted to be a risk factor for adolescent depression through learned hopelessness (27).

#### Sociocultural Contexts

The society, the physical environment and the culture in which the young people operate and live in was noted to play a significant role in how the youth experience depression. Firstly, culture determines how the youth and their families or society in general, define depression-the social interpretation and meaning of depression (25,27). In some cultures, within the SSA, someone who is depressed is considered to have deviated from the prescribed social norms (27). Others believe that depression is a cultural illness which happens due to witchcraft: “*I don’t know exactly if this is a cultural illness, the ‘snake in the stomach’, but it’s been reported to me probably four times in the time that I’ve worked in the township communities where those particular symptoms [of depression] have come out.”*(Community health worker, South Africa)(27). As already alluded to, culture influences how people in a particular society communicate their feelings and this is the basis of local idioms of depression or distress. Secondly, cultural philosophies and practices can either promote or demote mental health of young people. For example, the *Ubunthu*, a common Afro-cultural philosophy of ‘humanity’ which promotes peaceful co-existence, love, caring for one another and this in turn, promotes social cohesion and unity among community members (27): *“It means that you have to do onto me what you’d love done onto you”* (Social worker, South Africa) (27). With such a philosophy, the responsibility of taking care of children is not only with the biological parents but it becomes a shared responsibility among community members. Thus, burden of caring for a young person who is depressed is shared among the extended family and the immediate community members, at times (27). Additionally, the *ubuntu* practice acts as a defence mechanism against developing depression among members of the society, including the youth, especially in the context of maternal depression, through communal parenting (27). Thirdly and lastly, in SSA’s societies characterised by war, it was found that youth with depression have more externalising features of depression like irritability and antisocial personality traits like rudeness (25).

### 1. Experiences of Parents and Guardians of youth with Depression

The second objective of this review was to explore the experiences of parents and guardians of young people with depression. We found three main themes related to the experiences of the guardians, and these include (i) Challenges faced by the parents, (ii) Perception of youth depression and (iii) experience with the Healthcare System.

#### Challenges

While taking care of a young person with depression, guardians face numerous challenges which eventually impact on their mental wellbeing as well. The challenges include financial or economic burden (30). Parents are burdened with cost of transportation, medical scheme, school losses as some guardians must move their ward from one school to another due to teasing which was impacting on the academic performance of their child (30). A guardian narrates the financial challenges: “*My problem is that I do not have money; I cannot manage financially because my child’s depression is costing me lots of money.”* (Female Parent, South Africa) (30), and this was echoed by a young person: *“it is a life of struggle for us, the management is costing my mother the little grant she is getting from the government.”* (Male adolescent, South Africa) (30). Other challenges identified by our review include lack of awareness about mental health in general, stigma, psychological burden: *“we are suffering, not knowing what else we can do. I have exhausted all options. I started taking my daughter to traditional healers, pastors, and local counsellors on the street, but nothing seemed to help. She will be okay for one day and returns to her world the next day”* (caregiver 2, Tanzania) (28). Poor communication between the depressed youth and their guardians (19,27,30) was also identified as a challenge as such, some parents are not always aware of what their child is going through. One particular guardian from South Africa only knew about the challenges and the suffering that her daughter was experiencing when a HCW confined in her: *“I only realised when she attempted the first suicide, this time when she went to consult a psychologist, she told the psychologist how she attempted to kill herself and I was so shocked what if she killed herself..”* (Guardian-5, South Africa) (30); *“We hardly talk, which was really making her depression worse, but I now see that I am not doing enough”* (Guardian-8, South Africa). The poor communication was even worse in the context of adolescent perinatal depression and sometimes, this would culminate into client-parent physical fights or verbal fights. *“I usually fight with my child, you know these kids can be stubborn, you tell her to do this, she will not do it, so we fight. Her elder sister is not usually around, so am with this one, so we f ight a lot, we hardly talk, which was really making her depression worse but I now see that I am not doing enough.”* (Guardian-8, South Africa) (30).

#### Perceptions about Youth Depression

Because of lack of awareness, among other reasons, the attitude and perception of guardians about the youth with depression is mostly negative. Some parents blame themselves for their child’s depression thinking that as a parent, they may have failed in their parental duties especially in the context of adolescent perinatal depression (18). Additionally, one of the risk factors or cause of youth depression was poverty and this was mostly poverty in the household. So, some parents took the blame on themselves as being responsible for this, hence they themselves also experienced depression. The perception of the guardians is influenced by the nature of symptoms that their ward exhibits. For instance, the perception is worse when the youth exhibit more externalising symptoms of depression. Externalising symptoms like irritability, are deemed as bad while internalising behaviours, like social withdrawal, are favoured and viewed as good behaviours (27,28).

In Tanzania and South Africa, most parents and guardians, do not perceive depression as a health condition but rather, laziness. “*Her father said it was laziness because her walking changed; she walked slower than usual.” (*Guardian, Tanzania) (28). This wrong perception of some depressive symptoms often results in increased misunderstanding between some guardians and the youth (30). Furthermore, the misunderstanding of depressive symptoms is more obvious when the depression predominantly presents with somatic symptoms, with little cognitive or psychological symptoms, as is the case with youth depression (28): “*The problem was that she slept the whole day, crying and complaining about a headache and chest pain…They could not figure out her problem at the clinic …. I finally realized it was depression.”* (Caregiver 1, Tanzania) (28).

The negative perception by parents prevents them from accepting that their child is sick and requires mental health intervention. “*All she thought was that I am faking everything, especially when I self-harmed.”* (Female adolescent, South Africa) (30). This may have a bearing on the health-seeking behaviours of the guardians hence, contributing to prolonged duration without treatment. Linguistic complexities and lack of local terms to describe depression worsens the guardians’ lack of conceptualisation and understanding of depression as some guardians in SSA still think that depression is a condition for the western world (23,25,27,29).

#### Experience with the Healthcare System

With all the experience of trying to make sense of what their child is going through, some guardians are let down and frustrated by the healthcare system. The frustration is there because either the services do not meet their expectations, or they are not even available in the first place: “T*here is no treatment for stress and depression…or maybe I do not know if they are treatable, I think there is no help perhaps until one is crazy. They [HCW] only provide counselling on living with HIV, self-acceptance, and HIV medication usage”* (Caregiver, primary education) (28). The guardians’ expectation is that when they seek care, they should reach a safe haven, unfortunately, this is not always the case as mentioned by a guardian: *‘I do not just understand how they operated because I think they were against her [my daughter] like they were not treating her fine; they were disrespectful, because I used to tell them what was happening but they would not listen to me. It makes me feel so bad and sometimes, I will cry and eeeem what made me to cry was they were not listening to what I was telling them about my child[‘s] situation.”* (Parent, South Africa) (30). Guardians have a high expectation that the healthcare system will ably manage their child’s condition. However, they get frustrated when they do not immediately see positive changes in their child (30): “*She is getting worse, she cannot cope, she is always collapsing, is it obvious that nothing is working out fine? Her mood keeps changing like the depression is coming on her most times.”* (Parent, South Africa) (30).

## Discussion

The main objective of this review was to systematically synthesize qualitative studies on the lived experience of depression by the youths and their guardians in SSA and to aggregate the findings of the selected studies. Our Review resulted into 14 unique studies from 8 SSA countries, representing views of 610 young people (10 to 24 years; 72% females) with lived experience of depression, 297 caregivers and 70 healthcare workers. Findings on the lived experience of youths were aggregated into eleven categories which were further grouped into three main synthesised findings of making sense of depression, mental health systems and services and contextual factors as key issues encompassing YP’s lived experience. Regarding the experience of guardians, three key themes were identified including the challenges they face, their perception of youth depression and their experience in navigating through the healthcare system.

### The lived experiences of depression by the youth

#### Making Sense of Depression

Our review found that the youth undergo a phase where they seem “lost” and confused with their state of mental wellbeing and brain function as they try to make sense of what’s going on with them. Our findings are congruent with the findings by Mccnan et al, 2012 and Dundon, 2006 who noticed the same phenomena among YP from Australia, USA and Canada, of being confused in the process of trying to make sense of their struggle (13,33). As found by our review, during this phase of their depressive episode, the YP are overburdened by an emotional turmoil which pushes them to socially withdraw from their peers and family, to self-harm and in some cases, to use negative coping strategies like indulging into drug and substance abuse (13). Unfortunately, they withdraw from the same people who at some point, are required to notice and link them to professional support (33). Making sense of depression was characterised by the YP’s feelings of being disconnected from themselves. This was also observed by YP from Sweeden and USA (1,8) and Westberg et al, 2020 described this phenomenon as *’drifting’;* where the YP are observed to be “fumbling with life” due to the depressed episode (34). In our study, we found that the confusion which comes while making sense of depression, is exacerbated by the feelings of being different from others. Dundon 2006, talks of a similar situation where the perception of being different was associated with personality changes and associated loss of identity among American and Canadian youths (33). Furthermore, Dundon (2006) observed that the feeling of being different from others, as found by our review, especially among ALWH, makes depressed youths live in denial, believing that what they are experiencing is just a normal phase of life and not a medical condition.

In our review, we found that some of the depressive symptoms like ‘thinking a lot or deeply’, that the youths in SSA experience do not always conform with those listed in the DSM or ICD diagnostic criteria. This was also the case with some YP from the Western world (8). This is likely to result in a situation where some YP may be misdiagnosed as not being depressed when they are. Thus, a confusion arises when a YP considers themselves to be depressed while diagnostic tools are not able to pick that up (33). We thus, recommend the conceptualisation of depression to specifically fit the narrative of the youths. This conceptualisation comes in handy when we discuss the cultural influence on the experience of youth depression. This review observed that culture influences how a phenomenon like depression is experienced and communicated (35). We found that, the manifestation and interpretation of symptoms of depression vary even among SSA cultures. This has been observed by other studies as well, that in some cultures, somatic symptoms are more pronounced than in other cultures where the emphasis is on cognitive and/or emotional symptoms (36). Due to the differences in perception and expression of depressive symptoms as a result of cultural influences, diagnosing depression in some cultures is not easy. Furthermore, in some cultures, it is a taboo for people, especially, young people, to talk about their emotions (36). Linguistic complexities contribute to difficulties in communicating one’s emotions as most languages in SSA, do not have vocabulary for psychological disorders (37). Thus, this is likely to lead to cases of misdiagnosis and mismanagement. Additionally, our review found that, depression, as defined by the DSM or ICD, does not exist in some SSA cultures. However, in such contexts, there are local idioms of distress that resemble and share features with the DSM’s mood disorders of Depression and Anxiety. The most common local idiom of distress (IOD) that resembles Depression and is common in most SSA countries is ‘thinking deeply’ (38). Another common IOD is ‘having a heavy heart’ (38), which was described as ‘chest pain’ or ‘pain in the chest’ in our review. The Idiom of ‘having a heavy heart’ is synonymous with ‘thinking a lot’ in that, in some SSA cultures, it is believed that people think through their heart, hence, when they have a lot of thoughts in their heart, the heart becomes heavy (38). This resonates well with Proverbs Chapter 23 verse 7, which states that “For as he [man] thinketh in his heart, so is he…” (39). In conclusion, the findings of the experience of making sense of depression, which was associated with confusion, disparities in the symptoms et cetera reveals how youths from SSA and their counterparts from elsewhere, experience depression in a similar fashion with significant variations due to cultural differences.

Making sense of depression also entailed devising self-help strategies to control or minimize their distressing experience. Our review identified different strategies that are used by the youths in SSA; some of which were negative and counterproductive. A Systematic review and meta synthesis of how adolescents, majorly from High-income countries, experience depression found similar results (40). Like our review, they noted that YP use strategies like distraction, isolation and use of art and religion as a way of suppressing negative emotions (40). Interestingly, through our review and based on studies done elsewhere, we have noted that YP with depression almost use the same coping strategies regardless of the sociocultural contextual differences (40).

#### Mental Health System and Services

Exploring the lived experience of depression by the youth in SSA revealed how they navigate through the mental healthcare system in their respective countries. This includes the nature of the existing services, their experience in utilizing such services as well as the barriers they face while accessing such services. We identified services like screening and diagnostic services as well as treatment services like counselling. There is increased demand for more mental health services (MHS) by the youths (41). Thus, MHS are becoming more and more available than before (42). This is being necessitated by among other things, increased awareness about mental health, and increased availability and acceptability of Afrocentric psychological interventions like the Friendship Bench (43). Despite the numerous barriers to care that have been identified by this review and other articles outside this review (18,23,28,42,44,45), not all is lost in terms of youth mental health in the SSA region as the new (2025) Mental State of the World Report by The Global Mind Project indicates that African youths have higher levels of mind health and wellbeing compared to their counterparts from High-Income Countries, having scored highly on the Mind Health Quotient (MHQ) (46). The MHQ which measures a young person’s productivity and ability to navigate through life’s challenges, assesses a YP’s Mind Health and Wellbeing through the domains of Mood & Outlook, Social Self, Adaptability & Resilience, Drive & Motivation, Cognition and Mind-Body Connection (46).

#### Contextual Issues

Our review found that the context in which the YP experience depression shape and determine how they experience the depression. Three main contexts were identified. These include the HIV pandemic, Teen or Adolescent Pregnancy and Culture. HIV remains a significant public health burden in SSA with statistics indicating that 80% of all ALWH around the world are in SSA (47). It has been indicated that in 2022 alone, there were about 1.3 million new HIV case worldwide and. of these, 27% were young people aged between 15 and 24 years (48). Due to poverty, YP, especially girls, engage in multiple concurrent sexual relationships which predispose them to contracting HIV (48). Unfortunately, poverty is also a risk factor for youth depression (49,50). Hence poverty, which is rampant in SSA, predisposes YP to both Depression and HIV and the two conditions, as observed by our review, lead to a double burden of social stigma which is a perpetuating factor for depression among YPLWH (5,21). Due to the many challenges that YPLWH face, like issues around HIV Status disclosure, pill-burden and challenges with body image, when they are depressed, their experience is a complicated one, requiring extra support. Thus, most of the usual psychological interventions for depression may not fully work.

Teen pregnancy is another big problem of concern in SSA, where one-third of the teenagers become pregnant (51). The prevalence of Teen Pregnancy in SSA ranges from 24 to 30% (51,52). Factors associated with teen pregnancy in the region include early marriages, poor educational attainment, poverty, unemployment, lack of knowledge about contraception and unmet need for family planning (51,52). Teen or adolescent pregnancy is associated with increased risk for depression (53) with studies indicating a high prevalence of depression among pregnant teens of up to 40% (54). Suffice to mention that the relationship between depression and adolescent pregnancy is bidirectional with some studies indicating that in the presence of other risk factors, depression also contributes to early/teen/adolescent pregnancy (54). Depression among pregnant adolescent affects the outcome of the pregnancy in various ways including low birthweight (53).

Our review attested to the fact that the social-cultural context shapes the experience of a phenomenon. We noted how that culture influences how depression is viewed, defined, expressed and even experienced. For instance, our review found that in some cultures of SSA, depression is viewed as a cultural illness, associated with witchcraft. Similarly, some cultures in Australia, for example, view depression as an injury by the spirits (55). Some cultures in LMICs view depression as a “Rich people’s condition’ or a ‘condition of white people’ (55). While some cultural values and traditions are detrimental to the experience of depression, some cultures are protective and facilitate the healing process. Thus, culture contributes positively as well as negatively to the experience of depression depending on the context. Religion, as a system of beliefs, shapes the people’s culture in various ways and has been seen by some schools of thoughts as part of culture (56). It is an open secret that the majority (over 95%) of SSA people are religious with recent statistics indicating that slightly over 50% of people in Africa are Christians and 44% are Muslims (57). As observed by our review, some religious values, promote mental wellbeing and positive living. No wonder, some psychological interventions in Africa incorporate religious concepts like ‘Count your blessings’, which is practiced in Christianity, to help with cognitive restructuring for adolescents with depression (58). Systematic reviews have shown that Religion-adapted Cognitive Behavioural Therapy (R-CBT) effectively manages depression, and its use is gaining ground (59). In R-CBT, religious content and values are integrated into CBT to enhance cognitive restructuring and behavioural activation (59). Religion also determines how young people view and experience depression. For example, unlike in some western sectors where depression is viewed from the lenses of biomedical models, in most LMIC, it is viewed through other models like religion where, a depressed person is viewed as having a deficiency in faith in God (55).

### The Lived Experiences of Guardians of Depressed Youths

This review found that guardians face numerous challenges like financial burden when taking care of YP with depression. We found that these challenges eventually impact on the guardians’ mental wellbeing as well. Studies have indicated an increased risk of depression among parents taking care of children with mental health conditions like depression (60). For example, one study done in Ethiopia found that 58% of primary caregivers of children and adolescents with a mental disorder had depression (60). The duration of care and not being supported by other caregivers in the family, increased the risk of the depression (60). Unfortunately, when the guardians’ mental wellbeing is compromised, it backfires to the YP, and their depression worsens because, among other reasons, the guardians are less warm and less supportive (61). Overall, the quality of life for the guardians is compromised (62). A study investigating the quality of life (QoL) of caregivers of adolescents with depression in China found a decrease in the mean scores of both physical and mental QoL compared to the general population (63). This was associated with the course of the adolescent depression (AD), caregiver’s education and family functioning (63).

We further found that the majority guardians of YP with depression have negative attitude towards their children’s depression mainly due to lack of awareness and cultural values. We further found that some parents personalised their child’s suffering, viewing it as a failure on their part for failing to prevent the depression from occurring in the first place. This phenomenon unfortunately is not only found in SSA but also in other parts of the world. In the UK, for instance, it was found that guardians of YP with depression, commonly mothers, blame themselves for their ward’s depression, and in the process, they experience distress and uncertainty, they are helpless and are usually frustrated (64).

Furthermore, we found that due to lack of knowledge about depression, some guardians find it hard to identify symptoms of depression as real. For example, fatigue, a common symptom of youth depression, in the context of other symptoms like motivational anhedonia and social withdrawal was mistaken for laziness by some guardians in SSA countries of Tanzania and South Africa. This is congruent with a bottom-up review, co-written by experts by experience and academics, that in some contexts, depression is viewed as a personal incapacity and laziness (55).

Finally, our review found that guardians, generally do not have a pleasant experience when they decide to take their ward for professional care. Their expectations are not fully met by the healthcare system. They are thus, left frustrated in the process and end up going to informal healthcare pathways. This is not surprising as it has been indicated that just 0.62% of the health budget in SSA is allocated to mental healthcare (65). This means that services are likely to be scares, and when the services are available, they are likely to be unstructured, not youth-friendly (1) with corresponding high unmet needs (66).

## Conclusion

The Youths in SSA experience depression in a similar way as their counterparts from elsewhere with slight variations due to contextual factors like culture and medical conditions like HIV and Pregnancy, which are more prevalent in SSA. Unlike most systematic reviews that only focused on young people’s lived experience of depression, our review considered the experiences of their guardians as well. We have seen how that the guardians’ experiences shape the young people’s experiences of depression. The findings from this review will among other things, help mental health educators to contextualise their messages to suite the African narrative of Youth depression. The cultural variations of depressive symptoms will inform creation of diagnostic tools that are responsive to the cultural context of Africa. And finally, integration of African cultural and religious values may help to address the high unmet mental health needs among youths with depression.

## Limitations

The experiences reported in our Systematic review are not exhaustive and may not apply to all the youths with depression and their guardians in SSA. As such, they are not generalisable. However, they still give a picture of how youth depression is experienced in the context of the Sub-Saharan African. Researchers, Policy makers and even Practitioners may still use these results with caution.

## Other Information

### Registration and protocol

The review was registered with [PROSPERO](https://www.crd.york.ac.uk/PROSPERO/view/CRD42024556661) (CRD42024556661), where a brief copy of the protocol can be accessed. Additionally, the full protocol for the review was submitted to a journal and is under peer review. We report an amendment to the initial protocol on PROSPERO as follows: initially, we had planned to do a narrative synthesis using Framework analysis. However, after discussion amongst ourselves as researchers, we resorted to do meta-aggregation using the Joanna Briggs Institute (JBI) technique.

### Ethics approval and consent to participate

Ethics approval was not required for this systematic review as it only reviewed and reports on published studies that were duly approved by Institutional Review Bodies (IRB).

### Consent for publication

No additional Consent was sought as already available materials were included.

### Availability of Data and Materials

the availability of the dataset that will be generated during and analysed during this systematic review shall be reported accordingly after the systematic review is done. Suffice to mention that the data shall be found in the databases mentioned above.

## Data Availability

All relevant data are within the manuscript and its Supporting Information files.

## Acknowledgements

we acknowledge our two wonderful research assistants namely, Ken Rachid, Rabecca Kasale and Rachael Makanjira who assisted in study selection, data management and all administrative work related to the conduct of this systematic review.

## Availability of data, code and other materials

the following are available on request from the corresponding author: template data collection forms; data extracted from included studies; and analytic codes/ coding matrix.

